# A competency-based approach to pass/fail decisions in an objective structured clinical examination: An observational study

**DOI:** 10.1101/2020.03.24.20042093

**Authors:** Nazdar Ezzaddin Alkhateeb, Ali Al-Dabbagh, Yaseen Omar Mohammed, Mohammed Ibrahim

## Abstract

**Background:** Any high-stakes assessment that leads to an important decision requires careful consideration in determining whether a student passes or fails. This observational study conducted in Erbil, Iraq, in June 2018 proposes a defensible pass/fail decision based on the number of failed competencies.

**Methods:** Results were obtained for 150 medical students on their final objective structured clinical examination. Cutoff scores and pass/fail decisions were calculated using the modified Angoff, borderline, borderline-regression and holistic methods. The results were compared with each other and with a new competency method using Cohen’s kappa. Rasch analysis was used to compare the consistency of competency data with Rasch model estimates.

**Results:** The competency method resulted in 40 (26.7%) students failing, compared with 76 (50.6%), 37 (24.6%), 35 (23.3%) and 13 (8%) for the modified Angoff, borderline, borderline regression and holistic methods, respectively. The competency method demonstrated a sufficient degree of fit to the Rasch model (mean outfit and infit statistics of 0.961 and 0.960, respectively).

**Conclusions:** the competency method was more stringent in determining pass/fail, compared with other standard-setting methods, except for the modified Angoff method. The fit of competency data to the Rasch model provides evidence for the validity and reliability of pass/fail decisions.

## Introduction

The 21^st^ century has brought a worldwide shift towards a competency-based approach in undergraduate medical education ^1^. In medical education and assessment literature, ‘competence’ is the ability to perform a skill in a specific setting or situation, whereas ‘competency’ refers strictly to the skill itself ^2^. Adopting a competency-based approach to teaching necessitates changing the assessment methods^3^.

One of the greatest challenges to institutions responsible for training and certifying physicians is assessing clinical competence ^4–6^, which is significant because it helps to protect patients by determining whether candidates can progress to higher levels of study and/or medical qualification. Objective structured clinical examinations (OSCEs) are the most commonly used clinical competency assessment tools. If OSCEs are correctly designed and analysed, they can benefit medical students’ learning and future performance ^7^.

A key but difficult task in OSCEs is making pass/fail decisions in cases of borderline performance. To handle this issue, many standard-setting methods have been introduced and implemented in a range of clinical examinations ^8^. However, concerns regarding the reliability, validity and acceptability of these methods remain, particularly in the context of pass/fail decisions ^9^. The differences in cutoff scores among different standard-setting methods may reduce the legal defensibility of these cutoffs, especially when there are differences in the pass/fail decision ^10^. Pass/fail decisions are very important for students; failing an exam leads to remedial work, lost time and, under certain circumstances, the question of whether the student will continue or quit medical school^11^.

For standard setters, the presentation of performance data may contribute to the development of a credible and defensible pass/fail cutoff score ^12^. To recognise the relationship between the observed (actual) score on an examination and the underlying competence in the domain, which is commonly unobserved, a test theory model is necessary ^13^. Item response theory (IRT), which has received little attention in the medical education literature ^14^, provides a deeper analysis and gives a range of information on the behaviour of individual test items (difficulty), individual students (ability) and the underlying construct being examined ^13^. By using IRT models (e.g. the Rasch model), standard setters can identify items that are not mapped to student ability (i.e. items that are either too difficult or too easy for a particular cohort^15^. IRT models represent a powerful method for interrogating clinical assessment data, resulting in more valid measures of students’ clinical competencies to inform defensible and fair decisions on students’ progression and certification ^16^. Using Rasch measurement theory to establish competency level is a technique that can help decision makers by simplifying data in meaningful way and correcting for a mix of judge types ^17^.

Information gathered through assessment also directs students’ further formative development. Acknowledging this encourages transforming the assessment experience into a personal inquiry-based learning strategy ^18^ and listing potential types of learner deficits such as difficulties in clinical reasoning, history taking, physical examination or professionalism ^19^ to help set adaptive strategies.

The depth and breadth of research on OSCEs continue to expand, but a gap remains in the literature regarding pass/fail decisions in cases of borderline performance. This study aimed to examine pass/fail decisions using different standard-setting methods and to compare these with a new method based on the number of failed competencies. This will contribute to filling the knowledge gap regarding where to draw the line for the borderline performer and help to provide successful remediation. To achieve these aims, this study sought to answer the following questions:

Can the number of failed competencies facilitate pass/fail decisions? How accurately do the competency method data fit the Rasch model to provide a defensible pass/fail decision?

## Methods

This observational study was conducted in Erbil, Iraq, at the College of Medicine at Hawler Medical University.

### Participants

The data used in this study were examination results obtained from 150 final-year medical students completing an OSCE as part of their exit examination in June 2018.

### The OSCE

The OSCE included 23 work stations and two rest stations. A time of 5 minutes was allocated for each station, so each OSCE session took approximately 125 minutes. The examination was run in three parallel circuits on three different floors of the same hospital and was conducted in two rounds. Students were isolated between rounds to decrease the risk of sharing exam materials. Stations used real patients, trained simulated patients, manikins, or data and videos.

Each station’s score sheet contained a detailed checklist of the items examined. The checklist was scored with a maximum of 100 points. A global rating was also included for each station (1 = fail, 2 = borderline, 3 = pass, 4 = above the expected level).

### Validity and reliability

The content validity of the OSCE was established by using blueprinting to ensure an adequate sampling across subject areas and competencies, in terms of the number of stations covering each competency and the spread over the content of the tested course. The following skills were included in the blueprint as competencies ^20^: history taking (five stations), physical examination (six stations), data interpretation (five stations), procedural skills (two stations), communication skills (two stations—one station testing counselling skills and one history-taking station also assessing communication skills) and patient management (four stations).

For the quality assurance of the stations, question selection was conducted at both the department and the faculty level. The stations were prepared and reviewed for accuracy by the OSCE Committee, which included members from all departments. Stations were written in advance of the examination date, including instructions for the students, notes for the examiners, scenarios for the standardised patients, a list of requirements for each station and marking sheets. To ensure the consistency and fairness of the scores, training was conducted for both the examiners and the standardised patients.

Because each student taking the OSCE had to perform a number of different tasks at the stations, this wide sampling of cases and skills should result in a more reliable picture of a student’s overall competence. Moreover, as the students moved through the stations, they were examined by a number of different examiners, serving to reduce individual examiner bias.

### Standard-setting methods

For our examination of the exam results, standards were set using four different methods: a holistic score of 50% (university regulation), the modified Angoff (MA) method, the borderline regression (BLR) method and the borderline (BL) method.

Angoff standards^21,22^ for all stations were set by a group of eight experts. All experts had participated in teaching clinical sessions and assessing OSCE examinations. The experts were asked to define the criteria for a borderline (minimally competent) student. Through discussion, they reached consensus that a borderline candidate is a candidate performing at a level between ‘pass’ and ‘fail’. Using this definition, for each item on the checklist (on a scale of 0–100), the experts were asked to estimate the probability that a borderline student would perform that item correctly. For each item, experts differing more than 20% from the others were asked to discuss this with the other experts and to reconsider their judgment. The MA passing score was calculated for each station by averaging the estimates across experts and items. The MA passing score for the total exam was calculated by averaging the 23 station passing scores.

BLR ^23^ was the second method investigated. We performed a linear regression analysis, using student performance as total percentage scores and examiner global ratings (fail = 1, borderline = 2, pass = 3; and above the expected level = 4) to determine the cutoff score. The cutoff score was derived by substituting the borderline value (2) into the regression equation.

For the BL method, students with borderline performances were identified, and their checklist scores were collected. The mean score for this group was set as the passing score ^24^.

To assess the passing score for each competency, we first calculated the means of both checklist scores and global ratings of the stations assessing each individual competency, so each student had a mean checklist score and a mean global rating for each competency. The passing score of each competency was calculated using the BLR method, where students who failed more than half of the competencies (i.e. three or more of the six competencies) were determined to have failed the examination. Rasch item fit statistics were used to show how well the data for the competency method fit the Rasch measurement model. We then compared the pass/fail decision according to each standard-setting method with the pass/fail decision considering those who failed three or more competencies to have failed the exam.

### Data collection and data analysis

We did not require approval from our research ethics committee because this study was carried out using data acquired from normal exams in the curriculum (raw scores for each of the 23 stations for the 150 students in this study were obtained from the College of Medicine) with the goal of calculating the cutoff score for passing using different standard-setting methods and comparing these methods with the new competency method to improve student assessment and establish a defensible pass/fail decision. Thus, a retrospective analysis of existing data was conducted.

### Statistical analysis

SPSS, Version 23 and Excel 2010 were used for the data analysis. Cohen’s kappa was used to measure agreement between standard-setting methods on pass/fail decisions; this statistic can be interpreted as follows: Values ≤ 0 indicate no agreement, values of 0.01–0.20 indicate no agreement to slight agreement, values of 0.21–0.40 indicate fair agreement, values of 0.41–0.60 indicate moderate agreement, values of 0.61–0.80 indicate substantial agreement and values of 0.81–1.00 indicate almost perfect agreement ^25^.

### Analytical plan

A one-parameter (Rasch) IRT model was fitted to the data on 150 students and six competencies. We estimated competency difficulty based on how the student answered. Item difficulty is the value along the latent variables continuum at which a student has a 50% probability of passing each competency. In the Rasch model, the disparity between student ability and item difficulty predicts the likelihood of a correct answer. For example, if the difference between student ability and item difficulty is zero, there will be 50% likelihood of a student answering a question correctly. Higher item (competency) difficulty estimates indicate that students require a higher level of the ability to have a 50% probability of passing the competency ^14^.

An expected score will be calculated from each observed score in the Rasch process, using a *t*-test for each item. Item fitness to the Rasch model can be identified by means of infit and outfit statistics, which are expressed as ‘infit mean square’ or ‘infit *t*’ and ‘outfit mean square’ or ‘outfit *t*’. A value of 1 for outfit indicates a perfect fit, whereas values less than 0.70 indicate misfit and values greater than 1.30 indicate overfit. Infit *t* values also show the degree to which a question fits the Rasch model. Observed data follow the Rasch model if the results of infit *t* are non-significant (*t* from −2 to 2)^14^.

The item information function was calculated mathematically using the Rasch method by combining information on student ability and item difficulty. The sum over all items was plotted against student ability, giving the ‘test information function’ curve, which allowed the estimation of reliability at different levels of student ability. A tall narrow curve indicates a test containing highly discriminating items; less discriminating items provide less information but over a wider range ^14^.

## Results

The MA method yielded the highest mean passing score: 61.10 in the first round and 61.19 after discussion. This method resulted in a failure rate of 50.6%. The lowest cutoff score was produced by the holistic method, which had a failure rate of only 8%. The other standard-setting methods’ cutoff scores and failure rates are shown in Table 1.

**Table 1.** Standard-setting procedures applied to the 23 OSCE stations

| Standard-setting method | Passing score | Number of students failing | Failure rate (%) |
| --- | --- | --- | --- |
| MA | 61.19 | 76 | 50.6 |
| BL | 55.73 | 37 | 24.6 |
| BLR | 54.93 | 35 | 23.3 |
| Holistic method | 50 | 13 | 8 |
| Competency method | $\geq 3$ competencies | 40 | 26.7 |

OSCE: objective structured clinical examination; MA: modified Angoff; BL: borderline; BLR: borderline regression

To check for internal consistency reliability of the OSCE, Cronbach’s alpha was computed across the 23 stations for all students (n = 150) and was found to be 0.8.

Table 2 shows that failure in three or more competencies coincided with the student failure, as assessed by the holistic, BL and BLR methods, with 100%, 82.8% and 81% agreement, respectively.

**Table 2.** Percent agreement of the competency method with other standard-setting methods on which students failed the OSCE

| Number of failed competencies | Standard-setting method<br>N (%) |  |  |  |
| --- | --- | --- | --- | --- |
|  | MA | BL | BLR | Holistic |
| < 3 | 39 (51.3) | 7 (19) | 6 (17.2) | 0 (0) |
| $\geq 3$ | 37 (48.7) | 30 (81) | 29 (82.8) | 13 (100) |
| Total number of students failing | 76 | 37 | 35 | 13 |

OSCE: objective structured clinical examination; MA: modified Angoff; BL: borderline; BLR: borderline regression

Cohen’s kappa values for the different standard-setting methods on 3450 decisions across the 23 OSCE stations are shown in Table 3.

**Table 3.** Cohen’s kappa for the different standard-setting methods

| Standard-setting methods |  | Kappa | Sensitivity<br>(identified<br>a passing<br>student) | Specificity<br>(identified<br>a failing<br>student) | Mean kappa |
| --- | --- | --- | --- | --- | --- |
| MA | Holistic | 0.169 | 54 | 100 | 0.392 |
|  | BL | 0.483 | 65.5 | 100 |  |
|  | BLR | 0.457 | 64.3 | 100 |  |
|  | Competency | 0.457 | 64.9 | 94.9 |  |
|  |  |  | 62.175 | 98.727 |  |
| BL | Holistic | 0.449 | 82.5 | 100 | 0.653 |
|  | MA | 0.483 | 100 | 48.7 |  |
|  | BLR | 0.963 | 98.3 | 100 |  |
|  | Competency | 0.718 | 93.7 | 76.9 |  |
|  |  |  | 93.625 | 81.4 |  |
| BLR | Holistic | 0.475 | 83.9 | 100 | 0.652 |
|  | BL | 0.963 | 100 | 94.6 |  |
|  | MA | 0.457 | 100 | 46.1 |  |
|  | Competency | 0.713 | 94.6 | 74.4 |  |
|  |  |  | 94.625 | 78.775 |  |
| Holistic | MA | 0.169 | 100 | 17.1 | 0.379 |
|  | BL | 0.449 | 100 | 35.1 |  |
|  | BLR | 0.475 | 100 | 37.1 |  |
|  | Competency | 0.425 | 100 | 33.3 |  |
|  |  |  | 100 | 30.65 |  |
| Competency | Holistic | 0.425 | 81 | 100 | 0.578 |
|  | BL | 0.718 | 92 | 81.1 |  |
|  | BLR | 0.713 | 91.3 | 82.9 |  |
|  | MA | 0.457 | 97.3 | 48.7 |  |
|  |  |  | 90.4 | 78.175 |  |

OSCE: objective structured clinical examination; MA: modified Angoff; BL: borderline; BLR: borderline regression

Cohen’s kappa values ranged from 0.169 between the holistic and MA methods to 0.963 between the BL and BLR methods. The BL method had the highest mean kappa value (0.653), whereas the holistic method had the lowest mean kappa value (0.379). MA had the highest mean specificity to detect a failing student (98.727), whereas the holistic method had the lowest specificity (30.65).

The outfit and infit statistics showed that all competencies were within the acceptable range (both for mean square and *t* values) and accurately fit the Rasch measurement model (Table 4).

**Table 4.** Item difficulty, standard error, and outfit and infit statistics

| Competency | Item difficulty | Standard error | Outfit |  | Infit |  |
| --- | --- | --- | --- | --- | --- | --- |
|  |  |  | MNSQ | ZSTD | MNSQ | ZSTD |
| History | -1.05 | 0.2 | 0.919 | -0.51 | 0.937 | -0.54 |
| Examination | -1.78 | 0.27 | 0.82 | -0.62 | 0.847 | -0.95 |
| Management | -0.21 | 0.17 | 1.07 | 0.6 | 1.097 | 1.19 |
| Data | -0.52 | 0.18 | 0.706 | -2.84 | 0.754 | -2.99 |
| Skill | -1.3 | 0.22 | 1.198 | 1.09 | 1.094 | 0.77 |
| Communication | -0.76 | 0.19 | 1.048 | 0.43 | 1.041 | 0.45 |
| Discrimination | 1.31 | 0.16 |  |  |  |  |

Note: Item difficulty measured in logits (negative values indicate easier questions)
MNSQ: mean square (values ranging from 0.70 to 1.30 are within acceptable limits for the Rasch model); ZSTD: *t*-test (values ranging from -2 to +2 are within acceptable limits for the Rasch model)

Discrimination describes how well the OSCE items (competencies) separate students with abilities below the competency location from those with abilities above the competency location. One-parameter IRT often assumes fixed discrimination among all competency items. In practice, a high discrimination parameter (> 1) means that the probability of a correct response increases more rapidly as ability increases. Here, the discrimination value was 1.31, which indicates that the competencies better discriminate between high- and low-ability students than expected for items of this difficulty.

As shown in Figure 1, the competency of ‘examination’ was the least difficult, and ‘management’ was the most difficult. The change in difficulty shifts the item characteristic curves (ICCs), along with ability. The probability of success was higher for the competency of ‘examination’ than for the other competency items at any ability level. A student would only need an ability level greater than −1.78 on the competency of ‘examination’ to be expected to succeed on the competency.

**Figure 1.**
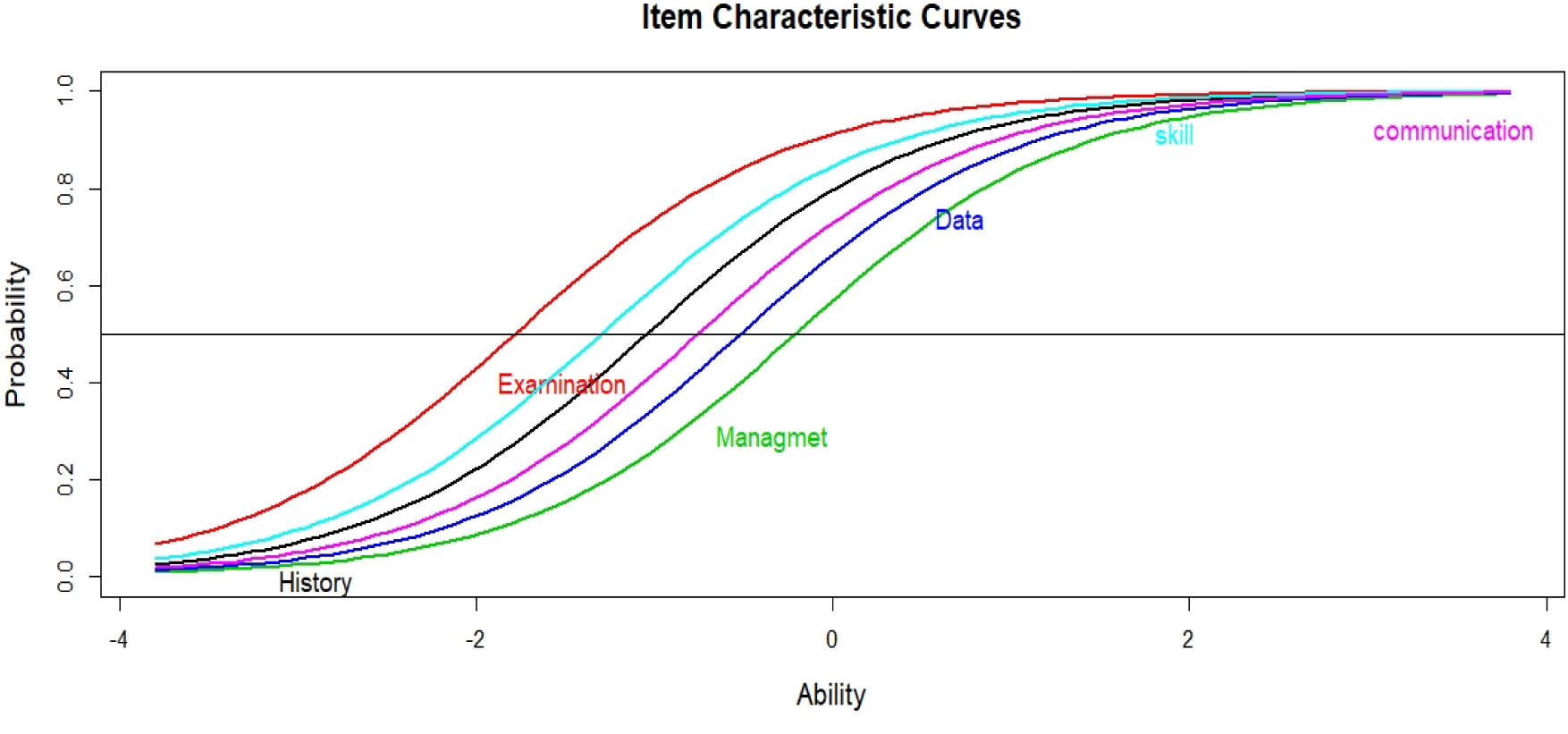
Plots of item characteristic curves for competencies in an item response theory model with competency difficulty levels of −4, 0 and 4. The competencies spread apart, representing varying levels of difficulty.

Similarly to the ICCs shown in Figure 1, the item information curves (IICs) shown in Figure 2 demonstrate that the ‘management’ item provides the most information about students’ high ability levels (the peak of its IIC is farthest to the right) and the ‘examination’ item provides the most information about students’ lower ability levels (the peak of its IIC is farthest to the left). All ICCs and IICs for the competencies have the same shape in the Rasch model (i.e. all competencies are equally good at providing information about ability).

**Figure 2.**
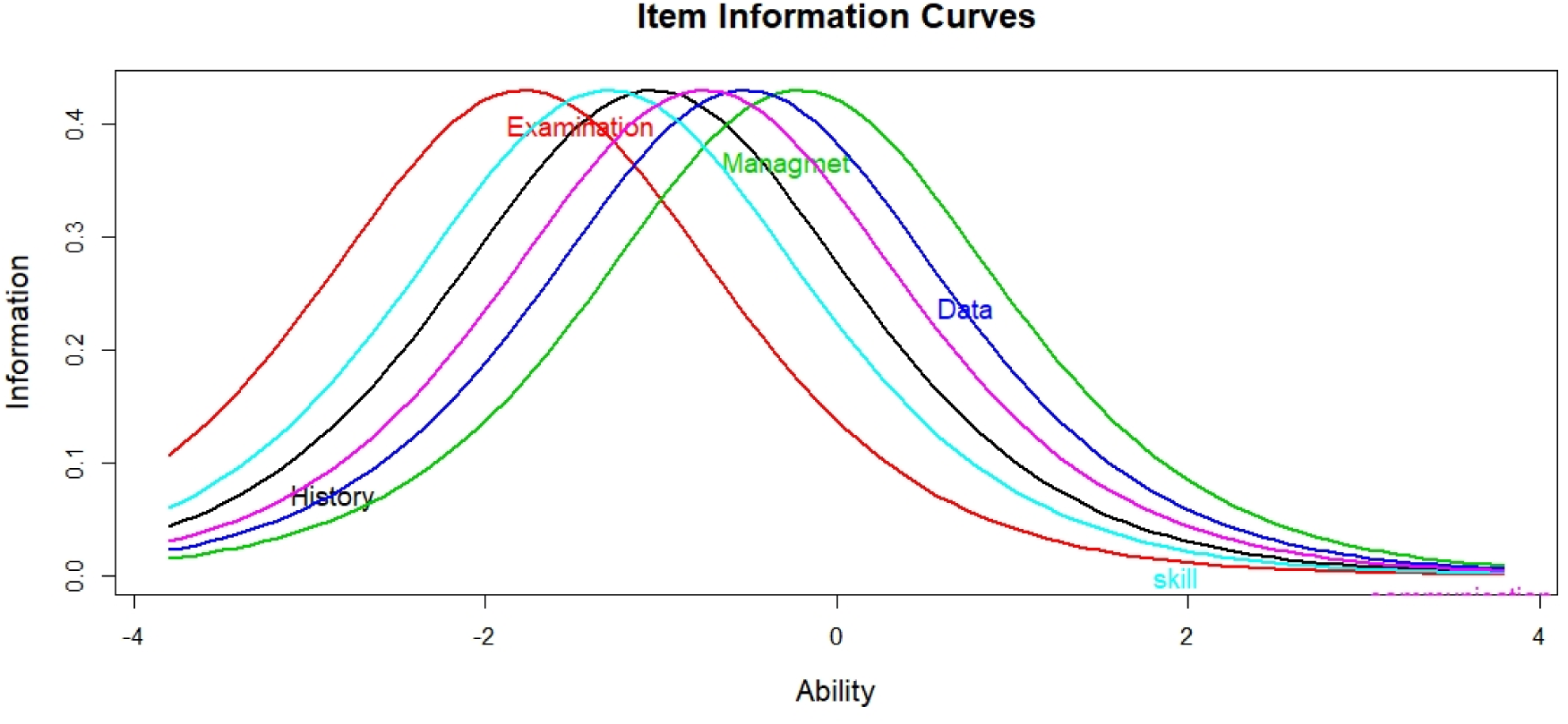
Plots of item information curves

The test information relied on the competencies used and the students’ ability. The test information can be calculated by summing all the competency information together. Figure 3 illustrates that the amount of information had a maximum at an ability level of approximately −1, when it is about 2.5. In other words, the competencies model is most informative when the ability of the student is equal to the difficulty of the competencies and becomes less informative as the student’s ability moves away from the competency difficulty (i.e. when the competency is either too easy or too difficult for the students).

**Figure 3.**
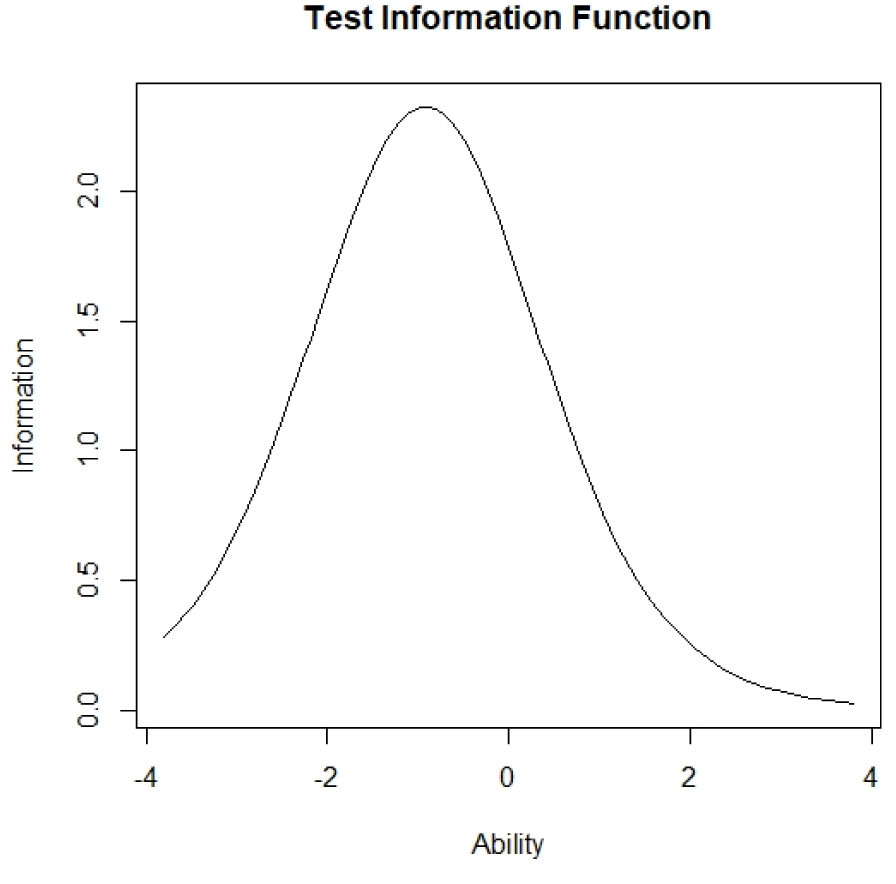
Item information function

## Discussion

The assurance of sufficient quality and robust standard setting is central to the delivery of any successful competency-based assessment ^26^. One of the most challenging aspects of clinical assessment is making pass/fail decisions for borderline grades allocated by examiners without adequate information to make these decisions ^27^. This study has proposed a new way of making pass/fail decisions in a high-stakes OSCE exam, incorporating the number of failed competencies, and examined the fitness of this new method to the Rasch model.

Different standard-setting methods may identify different cutoff scores for the same examination. Our study has shown diverging results, indicating there is no one valid ‘gold standard’ method. The ranking of the existing standard-setting methods from most to least rigorous is as follows: MA, BL, BLR and holistic methods. Despite the differences among standard-setting methods, the outcomes of the authentic methods (i.e. cutoff scores and pass/fail decisions) should be similar (if not the same) for the same examination ^28^.

Prior studies have addressed this issue and investigated the validity of different standard-setting methods for OSCEs ^28^. Lee et al. (2018) used the MA method to calculate cutoff scores for three different domains (history taking, physical examination and physician patient interaction). One major drawback of this approach is that the cutoff score for passing in the MA method (with reality check) increases or decreases when performance data are provided to standard setters ^12^. In the present study, we combined the OSCE station scores measuring the same competency, calculated the cutoff score for each competency and then we determined the pass/fail decision based on the number of failed competencies.

Different standard setting methods make different assumptions and determine cut scores differently. The competency method showed less agreement with the MA method, where only 48.7% of students who failed according to the MA method also failed when using the competency method. This difference might be caused by judges thinking about an average student instead of focusing on a borderline performer, leading to the substitution of a criterion-based concept with a norm-referenced one ^29^ and the setting of high cutoff scores. In contrast, the BL and BLR methods had the highest agreement with the other examined methods (mean kappa = 0.63 and 0.62, respectively). A previous study indicated that the BL and BLR methods produce more realistic cutoff scores, compared with the Angoff method ^30^. In the present study, the competency method has been shown to be more stringent than the other standard-setting methods except for the MA method, in terms of pass/fail decisions; using the competency method would therefore increase the number of students failing the OSCE. However, this result was expected and may be desirable because the negative consequences of certifying an incompetent examinee (false positive) may far outweigh those of not certifying a competent one (false negative). It is important to minimise passing incompetent students ^31^. According to course and programme leaders in a previous study, examiners in clinical examinations were too lenient and tended to avoid failing students, especially by giving borderline students the benefit of doubt ^27^. Such a practice has the potential to have major adverse implications for medical practice ^32^.

The second aim of the present study was to determine how accurately the data of the competency method fit the Rasch model. Classical test theory and IRT are widely used to address measurement-related issues that arise from commonly used assessments in medical education, including OSCEs. Traditionally, post-examination analysis results are often based on classical test theory. However, statistics in classical test theory are based on the aggregate, and their values are sample-size-dependent. Medical educators need to further investigate the relationship between students’ ability (independent of item sample size) and the ease or difficulty of questions (independent of student sample size). IRT and one of its main models (Rasch) offers a comprehensive and forensic analysis of exam data that can be used to enhance test quality ^14^. Furthermore, Rasch analysis provides beneficial graphical displays that aid test constructors in appraising the effectiveness of their assessments and, in the context of pass/fail decisions, enables us to establish the cutoff score for each competency according to student ability level. To judge the compatibility of the observed data with the Rasch model, mean square values were used: A value of 1 indicates a precise fit, whereas values from 0.70 to 1.30 indicate a good fit. However, values < 0.70 or > 1.30 are termed misfitting and overfitting, respectively, and should lead to an analysis of the items ^33^. Our results showed that the examined competencies accurately fit the Rasch model, with all competencies’ mean square values within the acceptable range. The Rasch analysis showed that the competency of ‘management’ was the most difficult, requiring greater student ability to pass.

In contrast to other standard-setting methods, which reduce all of the information obtained from an assessment to a binary pass/fail judgment and simplify high-stakes decision making such that a minimally competent student is treated the same as a maximally competent student—meaning that both can graduate as doctors ^34^, the competency method provides rich data on each student’s strengths and weaknesses. This presents an opportunity for students to learn from the assessment, guiding those who fail the examination in the remediation process and helping them to concentrate on their deficient competencies. The mantra that ‘assessment drives learning’ is often repeated with the belief that the effect of assessment is always useful ^34^. However, according to Kalet et al. (2012), effective remediation requires good data. In the present study, implementing the competency method in an OSCE exam provided data that could facilitate effective remediation.

The main strength of the present study is the comparison of many commonly used standard-setting methods for OSCEs. The reported Cohen’s kappa statistics were based on 3450 decisions over 23 stations. Another strength of the study is the use of the Rasch statistical IRT model to enhance the credibility of competency-based pass/fail decisions. Therefore, this new competency method is more dependable, as it is derived from mathematical principles, whereas other methods are based on an overall impression of the examination difficulty and provides a less defensible cut score.

However, obtaining data from final-year medical students in a single institution from one geographical region may limit the generalisability of our findings. It would be useful to include more students from different medical colleges in the region. Thus, our findings need to be interpreted with caution when applied to other institutional settings.

Our findings indicate the importance of combining the results of OSCEs based on content similarities of stations, which is more meaningful for a competency-based assessment and would enable faculty to draw more meaningful conclusions and provide actionable feedback.

Despite the implementation of many standard-setting methods in clinical examinations, concerns remain about the reliability of pass/fail decisions in high-stakes assessments, especially in the context of clinical assessment. The findings from this study make several contributions to the existing literature. First, the number of failed competencies help to establish a pass/fail decision, although this method is more stringent than other standard-setting methods examined except for MA. The competency method offers rich information about individual learners and provides data that facilitate effective remediation. The competency data fit the Rasch model well, which provides evidence for the validity and reliability of pass/fail decisions made using this method. The competency method can be used to set a cutoff score reflecting the desired student ability for each competency.

## Data Availability

Data will be provided on request

## Acknowledgement

We thank Jennifer Barrett, PhD, from Edanz Group (www.edanzediting.com/ac) for critically reviewing and editing a draft of this manuscript.

## Author contributions

NA, AA, MI designed the study, YM was the statistician and did the analysis. NA wrote the first draft of manuscript and data entry. All authors reviewed and approved the final manuscript.

